# The curvilinear relationship between the age of adults and their mental health in Iran after its peak of COVID-19 cases

**DOI:** 10.1101/2020.06.11.20128132

**Authors:** Jiyao Chen, Stephen X. Zhang, Yifei Wang, Asghar Afshar Jahanshahi, Maryam Mokhtari Dinani, Abbas Nazarian Madavani, Khaled Nawaser

## Abstract

The emerging body of research on the predictors of mental health in the COVID-19 pandemic has revealed contradictory findings, which prevent effective psychiatry screening for mental health assistance. This study aims to identify the predictors of nonsomatic pain, depression, anxiety, and distress, especially focusing on age as a nonlinear predictor. We conducted a survey of 474 adults in Iran during April 1–10, 2020, when Iran had just passed its first peak of the COVID-19 pandemic with new confirmed cases. We found that Age had a curvilinear relationship with nonsomatic pain, depression, and anxiety. Age was associated with pain, depression, and anxiety disorders negatively among adults younger than 45 years, but positively among seniors older than 70 years. Adults who were female, unsure about their chronic diseases, and exercised less per day were more likely to have mental health issues. This study advances the use of age as an effective predictor by uncovering a curvilinear relationship between individuals’ age and mental health issues by using a sample of adults across a wide spectrum of ages. We hope future research on mental health during COVID-19 pays more attention to nonlinear predictors.

## 1. Introduction

The COVID-19 pandemic has led to unprecedented disruption to people’s work and life (Zhang et al., 2020b) and triggered widespread mental health issues (Killgore et al., 2020; Xiang et al., 2020). A burgeoning body of research is unveiling the risk factors of mental disorders under the COVID-19 pandemic (i.e. Liu et al., 2020; Meng et al., 2020; Zhang et al., 2020b), however the evidence on individual demographics as predictors has been mixed. For example, some studies have found that age predicts mental disorders negatively (i.e. González-Sanguino et al., in press; Moccia et al., in press; Wang et al., 2020), but other studies have found the relationship between age and mental disorders to be positive (Qiu et al., 2020) or insignificant (Meng et al., 2020; Song et al., 2020; Zhang et al., 2020b). The conflicting evidence prevents the effective identification of the mentally vulnerable during the ongoing COVID-19 pandemic.

This study aims to investigate the role of age and other predictors on adults’ nonsomatic pain, depression, anxiety, and distress. In particular, this study does not assume the relationship between age and mental health disorders to be linear and instead examines its second-order term, using a sample of adults across a wide spectrum of age from 20 to 79. Furthermore, this study tests the predictors of individuals’ work situation, exercise hours, and the number of times out of home per day. Finally, following studies that found individuals among both healthcare workers and the general population who were unsure of their COVID-19 infection status had more mental health issues (Jahanshahi et al., 2020; Zhang et al., in press-b), this study tests individuals who were unsure of their chronic diseases as a predictor of mental health issues.

These predictors help to further advance the ongoing research on the predictors of mental health issues during the COVID-19 pandemic. The results provide some reconciliation of conflicting findings in this rapidly growing body of literature and provide some new directions for future research to specify risk factors and their boundary conditions to screen people in greater need to prioritize mental healthcare support.

## 2. Methods

### 2.1 Context

This study was conducted during the COVID-19 outbreak in Iran, one of the worst-hit countries early on. People in Iran faced some unique challenges such as US-led sanctions, an economic slowdown, a lack of medical equipment, and issues in enacting social distancing measures (Aman, 2020). People in Iran suffered from higher distress than people in China during the COVID-19 pandemic (Jahanshahi et al., 2020).

### 2.2 Data collection

Data in this cross-sectional study were collected through an online survey via social media platforms, e.g. Telegram, Instagram and WhatsApp, from April 1 to 10, 2020 in Iran. Iran passed the peak of its COVID-19 pandemic with 3,111 new infection cases on April 2 and had a total of 68,192 confirmed cases and 4,232 deaths of COVID-19 on April 10, 2020. The original survey in English was translated to the official language of Iran (Persian), pretested among 10 professionals in several industries, and approved by Shahid Rajaee Teacher Training University (IR.SSRI.REC.1389.685). Participation was fully voluntary and all participants agreed with their informed consent to complete the survey. We guaranteed the anonymity and strict confidentiality of responses and targeted working adults older than 18 years in Iran. Based on the number of active users in the social media groups, our survey could reach about 980 people. We received 474 usable responses, resulting in a response rate of 48.37%. The participants worked in banking, education, finance, insurance, manufacturing, services, mining, and oil and gas sectors from all 31 provinces of Iran.

### 2.3 Variables

Participants reported their demographic characteristics such as exact age, gender, marital status, and working situation. Participants also reported whether they had chronic diseases (no, unsure, yes), daily exercise hours, and the number of times they left home per day in the past week.

The outcome variables included nonsomatic pain, depression, anxiety, and distress. Nonsomatic pain was measured by a three-item including “For the past week I feel I am in agony” (0 = never, rarely, 3 = always; α = 0.75) (Keller and Nesse, 2006). Depression was measured by Patient Health Questionnaire-2 (PHQ-2) (0 = never, rarely, 3 = always; α = 0.77) with the cut-off point of 3. Anxiety was measured by the Generalized Anxiety Disorder-2 scale (GAD-2) (0 = never, rarely, 3 = always; α = 0.76) with the cut-off point of 3 (Kroenke et al., 2009). Distress was measured by K6, the six-item Kessler mental distress scale (0 = never, 4 = almost all of the time, α = 0.90) with the cut-off point of 13 (Kessler et al., 2002).

### 2.4 Data analysis approach

We used Stata 16.0 to summarize the variables and to predict pain by ordinary least squares regression and depression, anxiety, and distress by logistic regression with a 95% confidence level.

## 3. Results

### 3.1 Descriptive findings on the covariates

Table 1 shows that 37.4% of the 474 participants were younger than 40 years old, 50.2% were between 40 to 59, and 12.4% were above 60 years. Over half (51.3%) were female, and 87.1% did not have chronic diseases, 3.0% were unsure, and 9.9% had chronic diseases. Over half (56.3%) did not do any exercise in the last week, 37.3% exercised on average about one hour daily, and 6.4% exercised two hours or more per day. In work status, 44.9% worked at home in the past week, 26.8% worked in the office, and 28.3% did not work during the epidemic. 15.6% did not leave home in the last week, 37.8% left home once daily, and 46.6% left twice or more per day.

**Table 1.**
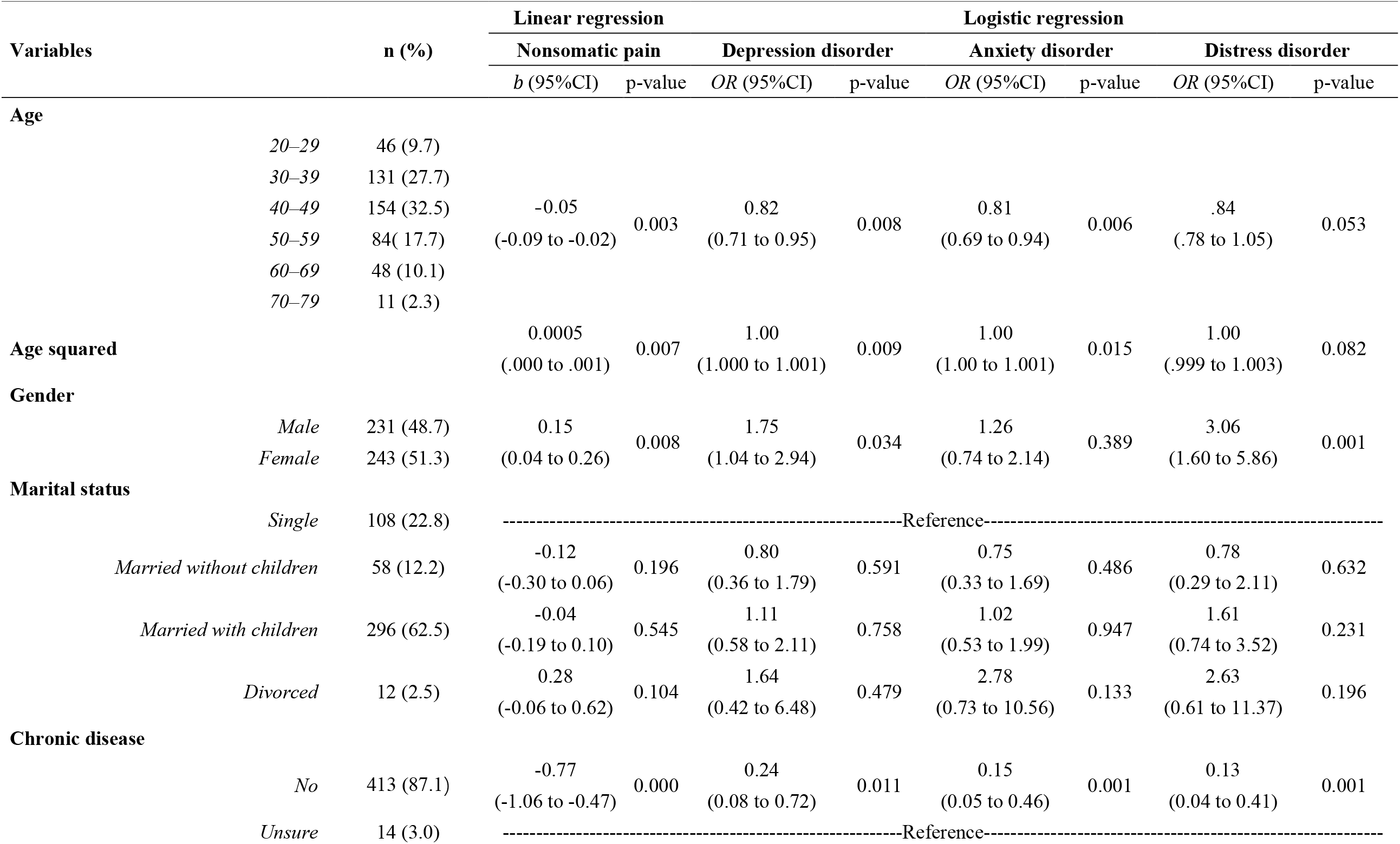
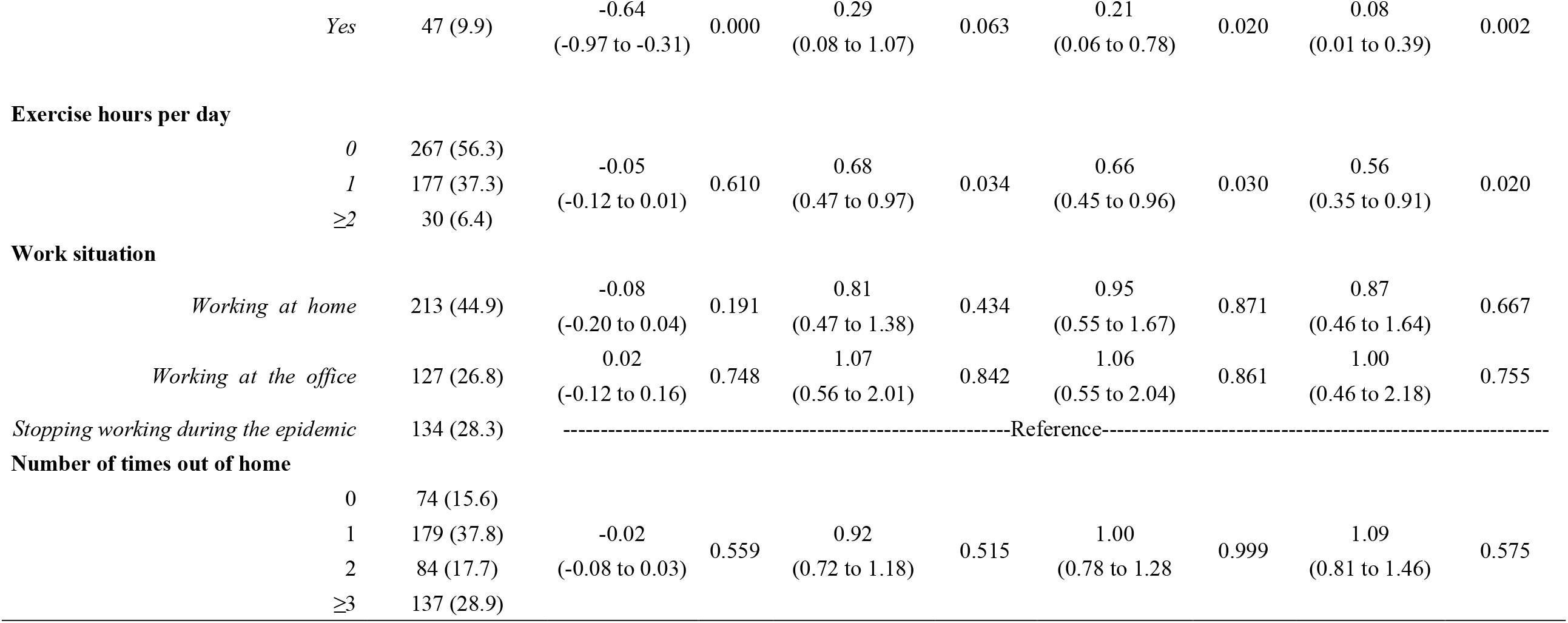
**The relationships between the covariates and pain (linear regression) and mental health disorders (logistic regression) (n=474)**

### 3.2 Descriptive and comparative findings on the outcome variables

Less than one in ten (8.65%) participants felt nonsomatic pain most of the time. About one fifth surpassed the cut-off of depression (21.94%), anxiety (21.10%), and distress (14.77%). By comparing our findings with those in 13 studies using similar measurements, we found that overall the mental health conditions of Iranian adults were comparable or worse than those in several samples in China, Spain, and Italy with a few exceptions (see Table 2 for a summary). For example, the proportion of adults with depression and anxiety disorders in our sample was similar to or worse than the samples from China and Spain during the COVID-19 outbreak with four exceptions. The adults in our sample were less likely to have depression and anxiety disorders than those in a sample of seniors older than 60 years in China (Meng et al., 2020) and of younger adults aged 18–30 years in the US (Liu et al., 2020), and to have depression than the adults in two relatively small samples in China (Guo et al., in press; Zhang et al., in press-a). The proportion of adults with distress disorder in our sample was also similar or worse than those in several samples in China and Italy, but lower than that in a sample of adults in the US in late April 2020 (Twenge and Joiner, 2020).

**Table 2.**
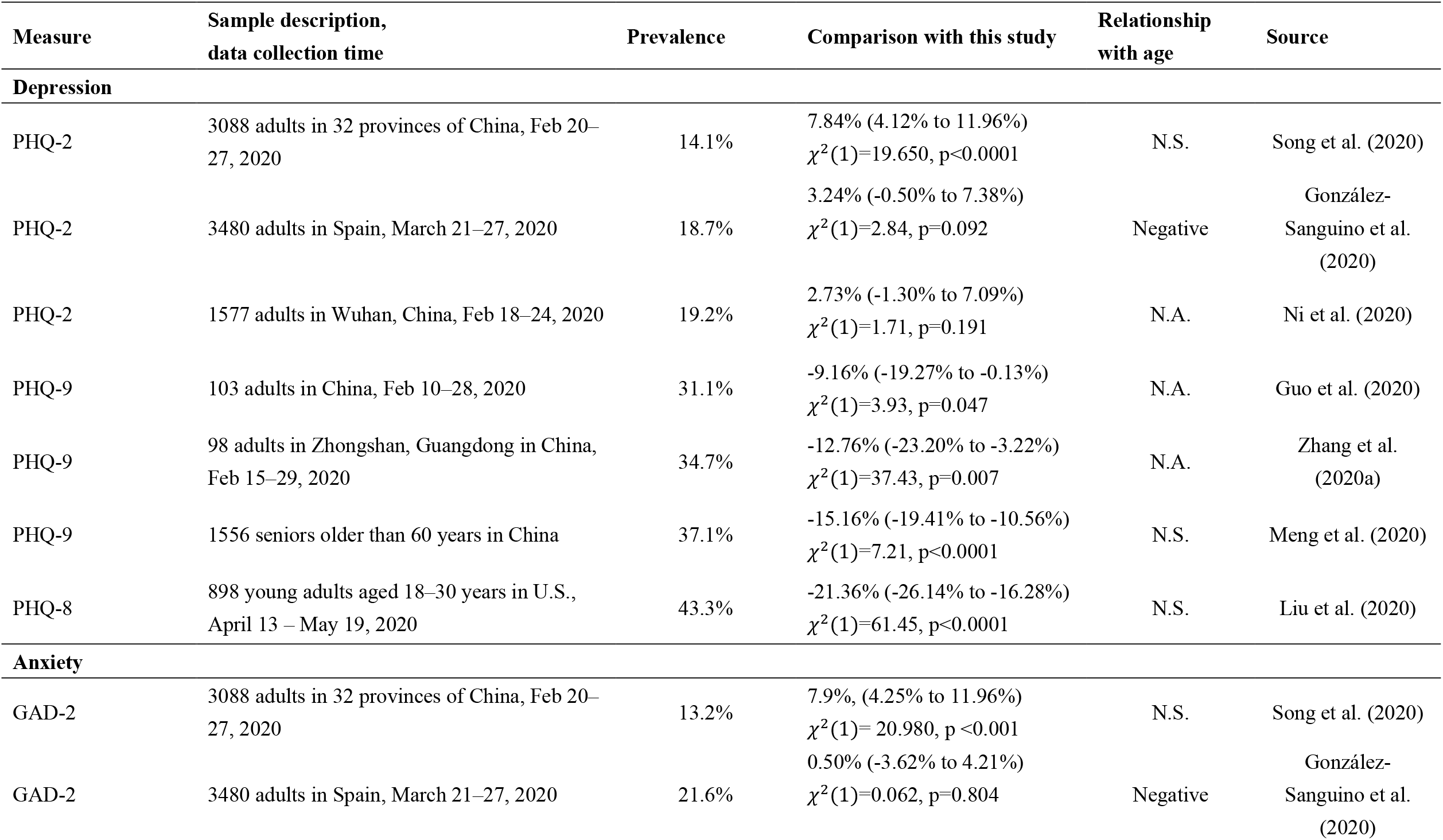
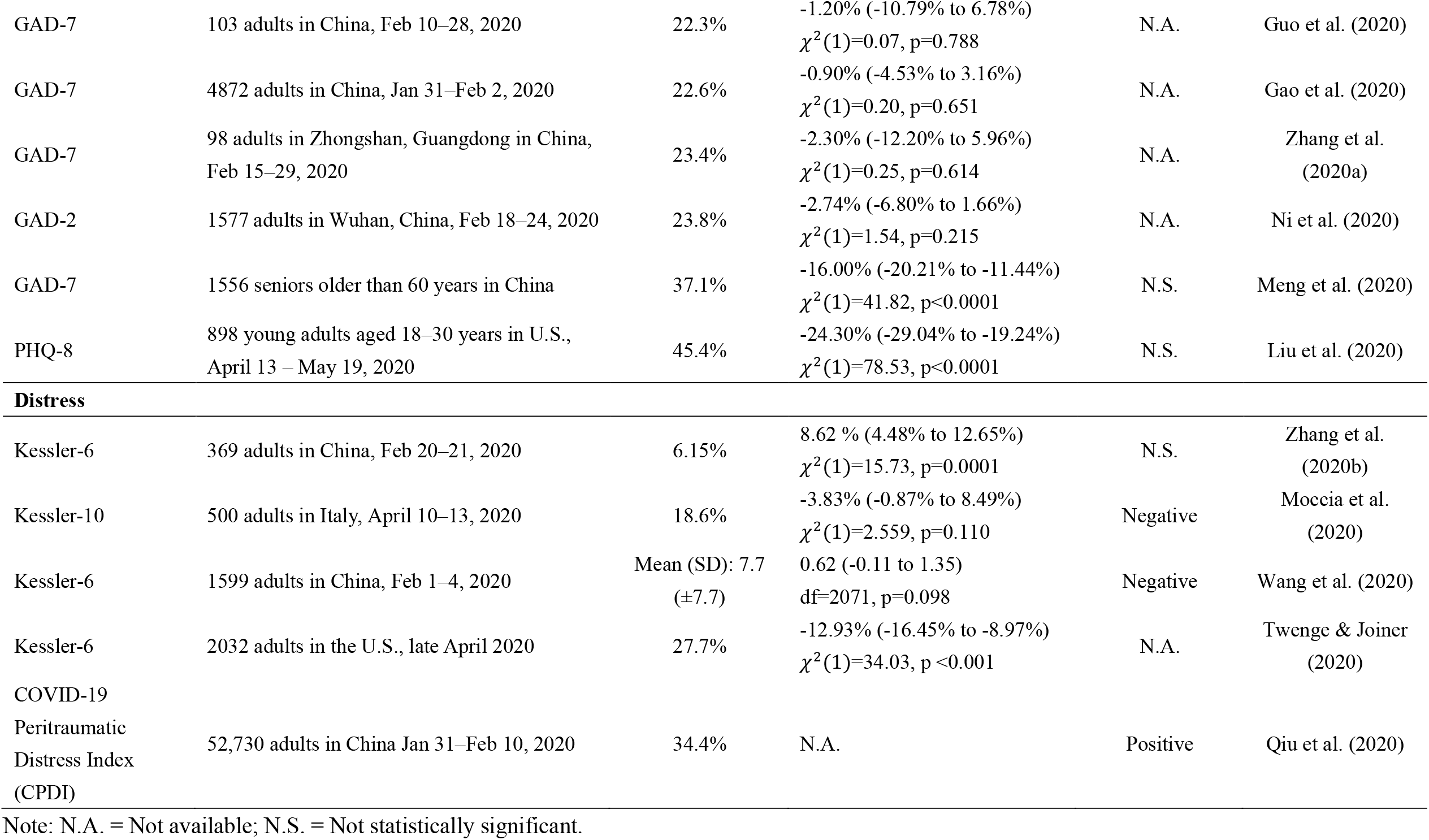
**The comparisons of adults’ depression, anxiety and distress issues during the COVID-19 pandemic across studies**

### 3.3 Predictors of pain, depression, anxiety and distress

First, adults’ age had a curvilinear relationship with their pain, depression, and anxiety (for pain: b=0.0005, 95% CI: 0.000 to 0.001, p=0.007; for depression: OR=1.00, 95% CI: 1.000 to 1.001, p=0.009; for anxiety: OR=1.00, 95% CI: 1.000 to 1.001, p=0.015). To show the curvilinear relationship, we plotted the mental health–age slope at varying ages in Figure 1. Margin effect analyses showed that adults’ age predicted their pain negatively among the young (e.g. at 20 years old: b=−0.032; 95% CI: −0.052 to −0.012; p=0.002). On the contrary, adults’ age predicted pain positively among seniors older than 70 years (e.g. at 75 years old: b=0.022; 95% CI: 0.002 to 0.042; p=0.030). Similarly, age predicted depression negatively among younger people (e.g. at 20 years old: OR=0.97; 95% CI: 0.95 to 0.99; p=0.006) and positively among older people (e.g. at 75 years old: OR=1.024, 95% CI: 1.01 to 1.05; p=0.042). Finally, age predicted anxiety negatively among younger people (e.g. at 20 years old: OR=0.97; 95% CI: 0.96 to 0.98; p=0.000) but not among older people (e.g. at 75 years old: OR=1.02; 95% CI: 0.99 to 1.04; p=0.214).

**Figure 1.**
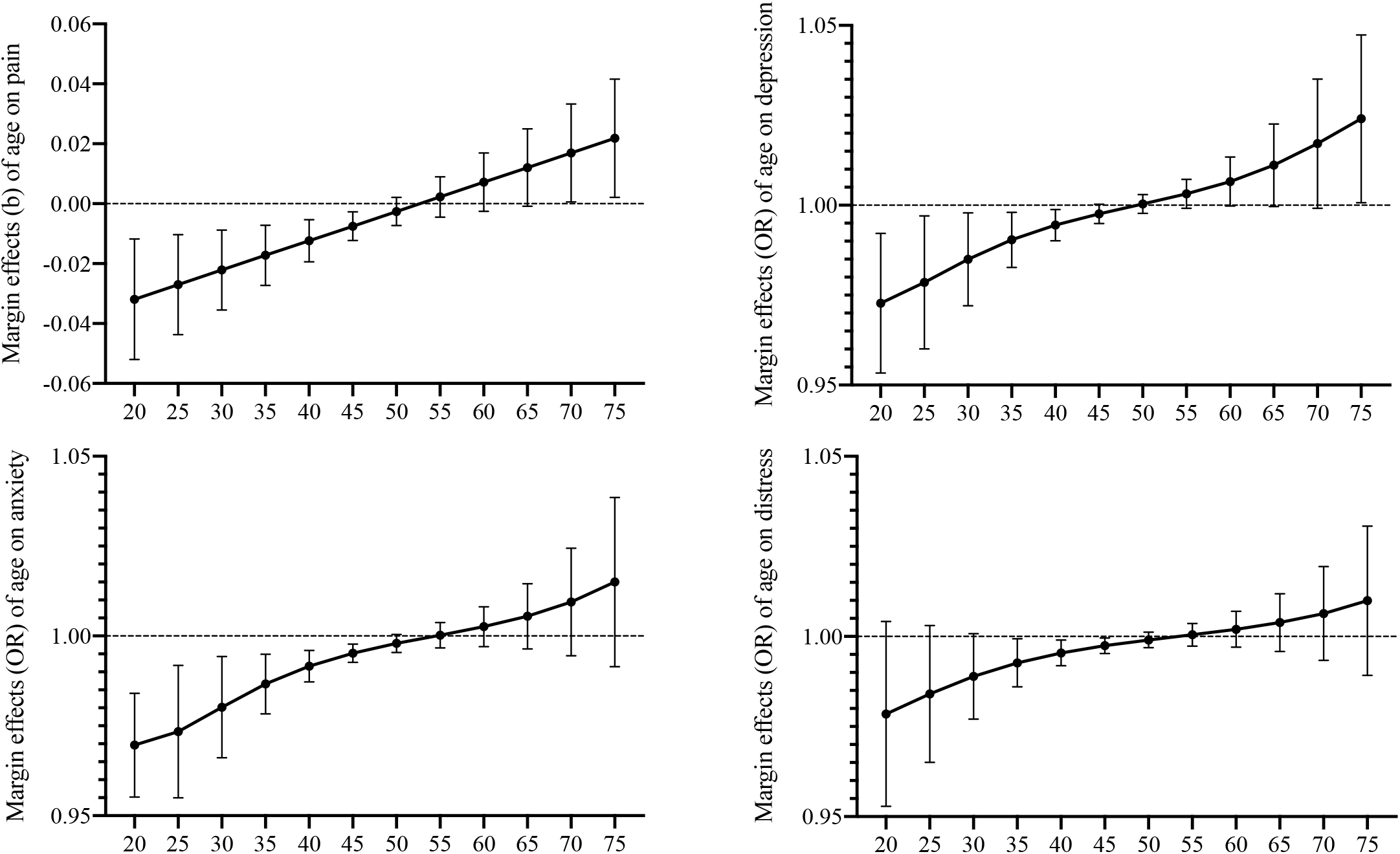
The relationship between age and (a) nonsomatic pain, (b) depression, (c) anxiety, and (d) distress disorders by age

**Figure 2.**
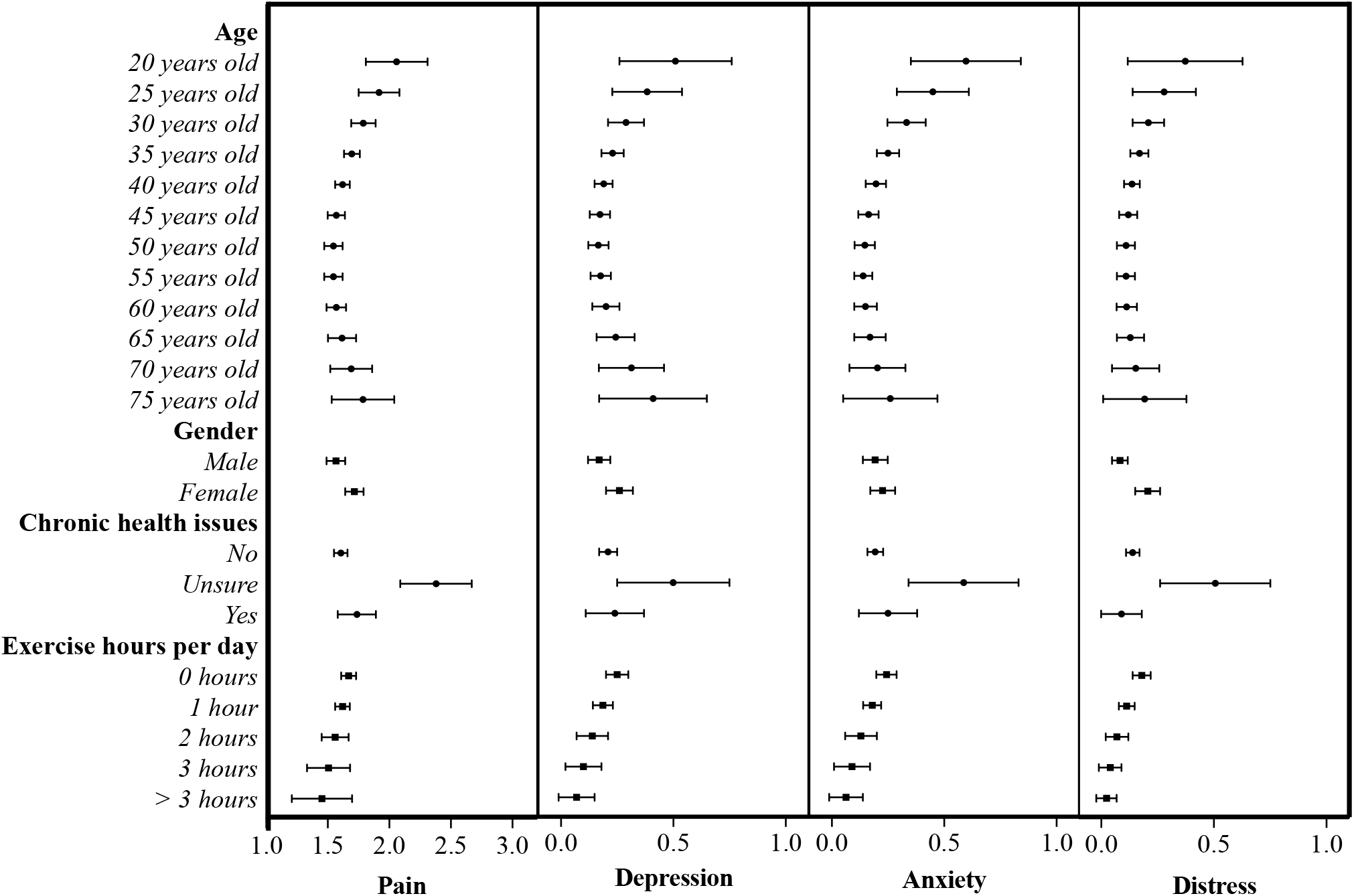
The predicted levels of nonsomatic pain and the predicted likelihood of depression, anxiety and distress disorders

Second, females reported more pain (b=0.15; 95% CI: 0.039 to 0.262; p=0.008), and were more likely to suffer from depression (OR=1.75; 95% CI: 1.04 to 2.94; p=0.034) and distress disorders (OR=3.06; 95% CI: 1.60 to 5.86; p=0.001) than males.

Third, adults who were unsure whether they had chronic diseases experienced more pain than those with (b=−0.64; 95% CI: −0.97 to −0.31; p=0.000) or without chronic diseases (b=−0.77; 95% CI: −1.06 to −0.47; p=0.000). Adults who were unsure whether they had chronic disease were also more likely to suffer from anxiety issues compared with those who with (OR=0.21; 95% CI: 0.06 to 0.80; p=0.020) or without (OR=0.15; 95% CI: 0.05 to 0.46; p=0.001) a chronic diseases. Furthermore, adults who were unsure whether they had chronic diseases were also more likely to have distress disorder, compared to those with (OR=0.08; 95% CI: 0.01 to 0.39; p=0.002) or without (OR=0.13; 95% CI: 0.04 to 0.41; p=0.001) chronic diseases. Lastly, those who were unsure of their chronic diseases were more likely to suffer from depression than those without chronic diseases (OR=0.24; 95% CI: 0.08 to 0.72; p=0.011).

Lastly, adults who exercised more were less likely to experience the mental health issues of depression (OR=0.68; 95% CI: 0.47 to 0.97; p=0.034), anxiety (OR=0.66; 95% CI: 0.45 to 0.96; p=0.030), or distress disorders (OR=0.56; 95% CI: 0.35 to 0.91; p=0.020).

## 4. Discussion

The accumulating body of studies on mental health under the COVID-19 pandemic has begun to show conflicting results, in particular on the relationship between age and mental health disorders (Table 2). Several studies found a negative relationship (i.e. González-Sanguino et al., in press; Moccia et al., in press; Wang et al., 2020); other studies found the relationship to be insignificant (i.e. Meng et al., 2020; Song et al., 2020; Zhang et al., 2020b) or positive (Qiu et al., 2020). The contradictory evidence may result from a) examining only a linear relationship and b) the limited ranges of age of the adults in the samples. Our study, with a wide range of ages from 20 to 79, showed that the relationship between age and pain, depression, and anxiety to be curvilinear (Figure 1). In particular, age predicted pain, depression, and anxiety disorders negatively among adults younger than 45 years. Age was not a predictor of pain for ages 50–65 years, for depression for ages 45–70 years, or for anxiety for ages 50–80 years. Age predicted pain positively among those aged 70–79 years and depression disorder among those aged 75–79 years. Such findings corroborate that the association between age and mental issues is negative among the younger population (< 45 years), insignificant among the mid-aged (50–70ish), and positive among seniors over 70 years. Seniors experienced more pain and depression, perhaps because they have a higher fatality rate during the COVID-19 outbreak and often have poor physical conditions and immune systems (Meng et al., 2020). In fact, a senior-older-than-60-year study (Meng et al., 2020) and a young-adult between-18-to-30 study (Liu et al., 2020) showed that they were much more likely to suffer from depression disorder (37.1% and 43.3%, respectively) than adults with varied ages in the other seven studies (on average 20.9%) (Table 2).

Hence, our results suggest age remains a useful predictor of mental health disorders during the COVID-19 pandemic, yet psychiatrists and mental health services need to be careful using age as either a positive or a negative predictor alone in identifying and screening people at mental health risks from the COVID-19 pandemic. Rather, they should be mindful that the association between age and mental health issues could vary depending on the age range. The boundary condition of age as a positive or negative predictor needs to be established before it can be used properly to identify the mentally vulnerable.

Our results showed that those who were unsure about their chronic diseases experienced more pain and were more likely to have depression, anxiety, and distress disorders than those who were sure about their status (either having or not having chronic diseases). Such a pattern was consistent with the findings that healthcare workers (Zhang et al., in press-b) and the general population (Jahanshahi et al., 2020) who were not sure whether they were infected by the COVID-19 virus were more likely to have mental disorders than others who did know their status. In doing so, we extend the finding from COVID-19 to chronic diseases, and suggest future studies explore individuals’ uncertainty on key factors as potential predictors of mental health.

Unlike past research that found gender was not a predictor of distress in China (Zhang et al., 2020b) and Iran (Jahanshahi et al., 2020), our Iranian sample showed that gender predicted distress, as in a Italian sample (Moccia et al., in press). In contrast to a study which found Chinese working adults who exercised less were happier with their life in a more severe COVID-19 prefecture (Zhang et al., 2020b), our results that Iranian working adults who exercised less were more likely to be depressed, anxious, and distressed corroborated the findings in a study of healthcare staff in Bolivia (Zhang et al., 2020a). All studies confirmed exercise time as a risk factor, but the direction of its effect may depend on countries and type of professionals. It is also worth noting that, in contrast to adults in China (Zhang et al., 2020b) and in Iran before the peak of COVID-19 (Jahanshahi et al., 2020), those who stopped working were insignificantly different from either those who worked at home or at the office on reported pain, depression, anxiety and distress. These studies show the importance of the boundary conditions of gender, exercise time, and working status in screening mentally vulnerable people in the crisis. The inconsistent findings call for more studies to investigate these predictors of mental health issues and their potential moderators such as country and stage of the pandemic.

This study has some limitations. First, due to the challenge of data collection during the COVID-19 crisis, we used convenient sampling; future studies with more representative sampling techniques could further examine age as a predictor of mental health. Second, the curvilinear relationship that we found was limited to our sample in Iran, and it is interesting to examine how it may vary in other populations. Nonetheless, our results point to the need for further studies to determine how age plays out as a predictor to enable better identification of those who are in greater need of mental healthcare.

Our study showed that Iranian adults after the peak of COVID-19 still suffered from depression, anxiety, and distress disorders to a similar or higher degree than people in other countries, except the US. The finding that the association between age and mental health issues was curvilinear suggests age remains a useful yet more nuanced predictor than deemed by the past literature. As the research on mental health under COVID-19 progresses, we call for more studies to examine the nonlinear relationships of the predictors of mental health issues.

## Data Availability

The data used to support the findings of this study are available from the corresponding author upon request.

## Acknowledgement

Nil.

